# What may work in deprescribing? A scoping review of intervention types, targets and outcomes

**DOI:** 10.1101/2025.07.30.25328075

**Authors:** Eabha Manley, Zeynep Or, Matthias Brunn

**Affiliations:** IRDES, EHESP; IRDES; Sciences Po – LIEPP

## Abstract

**Background:** The overuse of medications, especially in older populations, is a growing public health concern. Not only does inappropriate prescribing contribute to adverse drug reactions, but it also results in significant economic and environmental impacts. The process of deprescribing, or the systematic reduction or cessation of medications, aims to improve health outcomes and address the risks associated with polypharmacy.

**Objective:** This scoping review seeks to map the various types of deprescribing interventions, their target populations, and the outcomes associated with these interventions. It provides an initial assessment of their effectiveness and synthesizes evidence from multiple disciplines to inform broader public health strategies.

**Methods:** A comprehensive literature search was conducted using PubMed and Web of Science to identify studies from 2010 onward that evaluated deprescribing interventions. The review includes interventions targeting different patient populations, healthcare settings, and medication types, with data extracted on intervention characteristics and outcomes.

**Results:** Of the 179 studies included, the majority focused on older adults, with a significant number targeting polypharmacy. Most interventions involved provider-focused strategies, including education and medication reviews, with some also engaging patients directly through educational materials. Pharmacists played a key role in many effective interventions. While outcomes varied, interventions consistently demonstrated positive impacts on medication reduction and patient safety, with minimal adverse effects.

**Conclusion:** This review highlights the importance of multidisciplinary approaches, patient involvement, and guideline-based tools in effective deprescribing. The findings support the implementation of deprescribing interventions as a public health strategy to reduce medication-related harm and contribute to environmental sustainability. However, further research is needed to assess long-term efficacy, the impact on younger populations, and the environmental outcomes of deprescribing.

## Introduction

Drug consumption is a key feature of healthcare systems and increasing globally. Between 2010 and 2022 spending on medicines has grown by almost 70%, reaching 1.48 trillion US dollars worldwide. At the same time, a considerable proportion of prescriptions are deemed inappropriate. More than half of older people were prescribed at least five drugs in most European countries which increases the risk of adverse drug reactions.^1^ In France, an estimated 54% of nursing home residents and 25% of community-dwelling older adults receive potentially inappropriate prescriptions^2^, contributing to a 136% rise in hospital admissions for adverse drug reactions between 2007 and 2018^3^.In primary care, the UK’s National Overprescribing Review estimates that at least 10% of the total number of prescription items need not have been issued.^4^

The over- and inappropriate consumption of medicines not only compromise the quality of care and escalate healthcare costs, but also exert a substantial environmental burden. In 2018, French households discarded about 17,600 tonnes of unused medications, contributing to water pollution that threatens aquatic life and fosters emergence of resistant bacteria.^5^ Moreover, pharmaceuticals are among the largest sources of CO2 emissions within the health sector, representing about 25% of healthcare related GHG emissions in Canada and 33% in France.^6,7^ These figures illustrate the major environmental and public health impacts of pharmaceutical overconsumption. Therefore, transforming drug prescription and consumption patterns to minimize waste and inappropriate utilisation is a critical public health priority.

Deprescribing is defined as the planned and supervised process of reducing the dose or stopping the use of one or more medicines, with the aim of improving patients’ health outcomes. It involves a careful assessment of the benefits and risks of each drug prescribed, to identify inappropriate or low value prescriptions to develop a plan to stop their use.^8^ Despite its potential benefits, deprescribing medicines is challenging and faces several barriers. These include the need for consultation between different healthcare providers often with conflicting incentives, resistance from patients, restrictive regulatory frameworks, and the pressure from pharmaceutical industry. Lack of coordination and communication among stakeholders can obstruct the implementation of effective deprescribing interventions, reducing their capacity to consistently minimize inappropriate medication use.^9,10^

Current research on deprescribing intervention generally focuses on certain population groups and intervention types, such as pharmacist-led interventions^11^, benzodiazepine use^12^ and elderly patients.^13^ Yet, there is a lack of a broad perspective on deprescribing strategies across different populations and care settings. This scoping review aims to map deprescribing interventions, their target populations, and associated outcomes, providing an initial assessment of their effectiveness. By synthesizing evidence from a wide range of domains this review broadens the discussion to include public health disciplines like policy, management, economics, and epidemiology. The question “what works in deprescribing?” seeks to guide practitioners, researchers, and policymakers in translating the evidence into scalable, impactful public health policies.

## Methods

Considering the variety and volume of evidence available on deprescribing we carried out a scoping review to identify major types of interventions, their target populations, and outcomes. We used the methodology developed by Arksey and O’Malley for scoping reviews^14^ and PRISMA-ScR, the PRISMA extension for scoping reviews in reporting.^15^

### Search strategy

The search strategy was developed and conducted in collaboration with a research librarian. We searched PubMed and Web of Science using a list of keywords in varying combination that was developed and discussed by the research team (including deprescription, deprescribing, polypharmacy, interventions, medication, discontinu*, policy) (see Annex 1). The initial database search was carried out on 17/4/24 with additional searches performed on 19/6/24 and 25/7/24. Following our search, we used snowballing to identify additional articles by reading reference lists of published reviews that were identified in our search.

### Eligibility criteria

We included English or French language studies performed in any country that described and evaluated a deprescribing intervention. An intervention is a clearly defined and coherent program or policy, or a specific and clearly defined practice, as opposed to usual care in a system or a one-off activity in a drug trial. We targeted interventions that specifically aimed to stop or reduce medication use rather than improving the quality of prescriptions. We did not place any restrictions on population characteristics or study setting. We only included studies that evaluated the impact of interventions using any type of outcome and included articles from 2010 onwards. We excluded opinion pieces, qualitative studies, study protocols of unpublished trials, literature reviews, and grey literature.

### Study selection

All titles and abstracts from our literature search were reviewed for inclusion by 2 members of the research team. Where there was discordance amongst team members regarding inclusion, the title and abstract were reviewed by a third team member to reach consensus. Full texts of these articles were then reviewed for inclusion in the final study. When a paper was excluded, a reason was recorded for explanation.

### Data extracting

To extract data from selected articles, the following categories were identified collaboratively by the research team: study type, intervention setting, population targeted, classes of medications targeted, details of the intervention (including intervention content and intensity, disciplines involved, target of the intervention, guidelines or tools used, and outcomes based on the evaluation of the intervention. We applied the GRADE^16^ criteria to assess the quality of the included studies assisted by ChatGPT version 4. One author reviewed all ratings for consistency, while two additional authors randomly checked a subset of 10 papers each. All data was then tabulated and analyzed to identify patterns.

### Data availability

Our full data extraction sheet is available as supplementary material.

## Results

Our initial database search produced 596 articles of which 150 were duplicates. A total of 446 titles and abstracts were reviewed for inclusion following which 260 were selected for full text analysis. Four full text articles were not available and 77 were deemed unsuitable due to not being deprescribing specific or not assessing an intervention. Eventually, 179 articles were included in our review (Figure 1).

**Figure 1.**
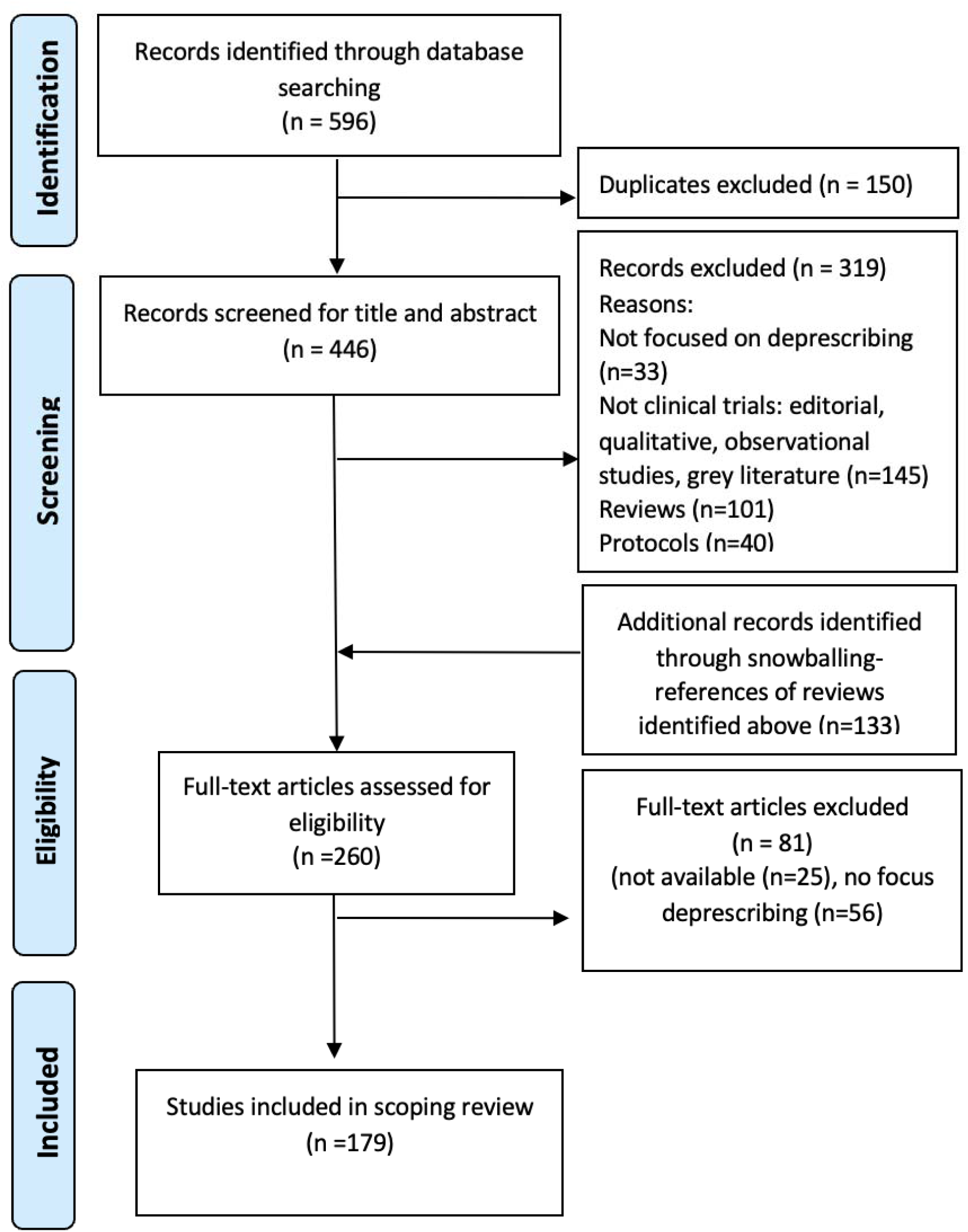
Flowchart of the study selection process.

### Geographic origin and year of publication

The USA had the highest number of publications with 42 studies, followed by Canada with 34 studies: four from the D-PRESCRIBE trial cluster, three from the MEDSAFER trial, and two from the EMPOWER cluster. Ten studies took place in Australia and the rest came from Europe, South America, Asia and the Middle East. Of 179 articles, 71% were published after 2017 and 44% were published after 2020 (Figure 2).

**Figure 2.**
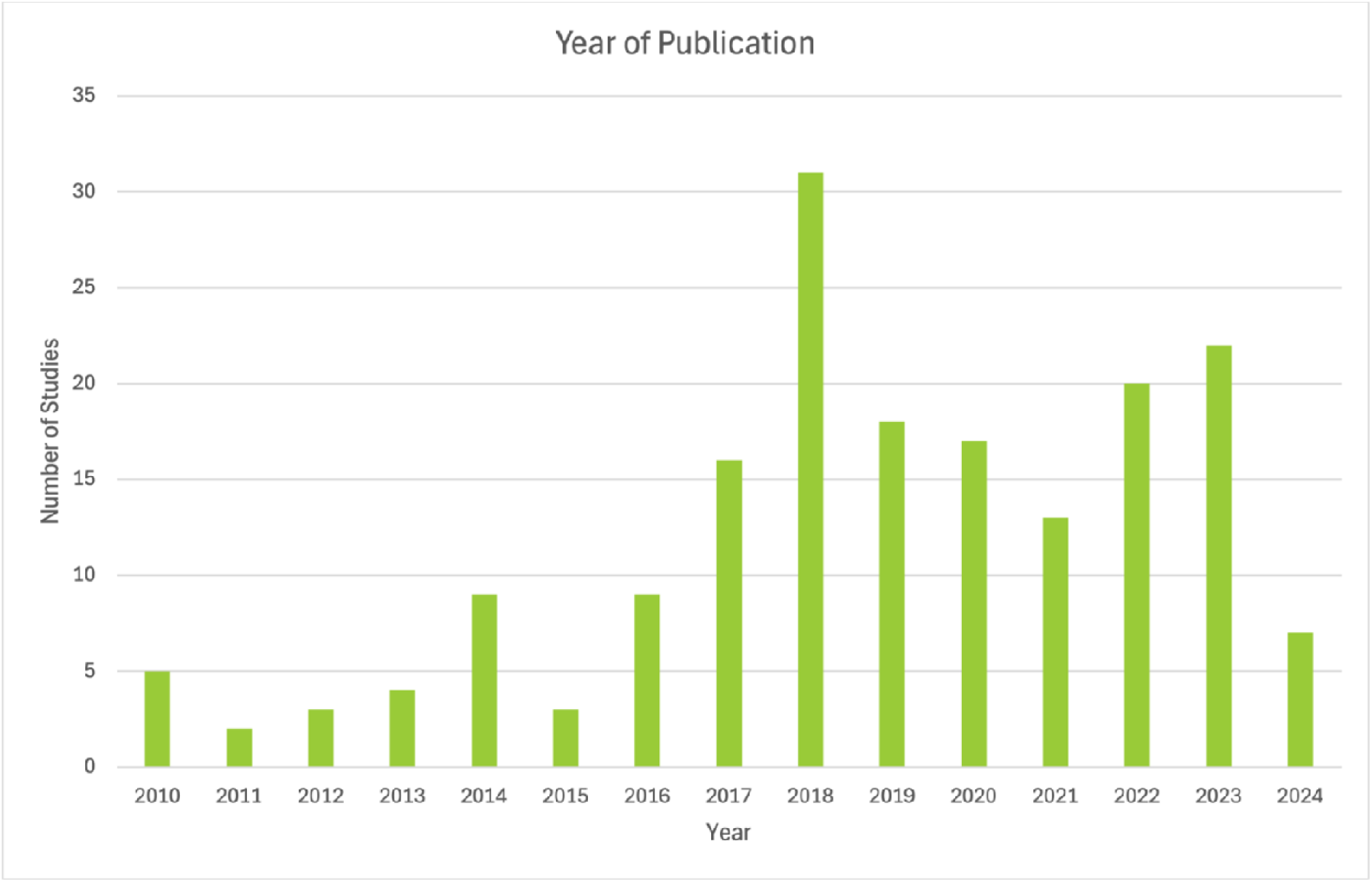
Year of Publication.

### Study type

We classified studies into one of the following four groups: RCTs, quasi-experimental studies, observational pre/post studies, and pilot or feasibility studies. 70 studies were RCTs, of which 21 were cluster RCTs. 12 studies were quasi-experimental, 40 were pre-post interventional studies, and 57 were pilot or feasibility studies. 80% of RCTs compared interventions to usual care. Sample size ranged from 10 participants to 163,776, with a median sample size of 190 participants.

### Setting

The majority of interventions (53%) took place in community/outpatient settings, including one in an outpatient dialysis unit and two in specialist mental health outpatient clinics. 21% of studies reported on interventions in inpatient setting, including one in an intensive care unit. 20% of the interventions were carried out in nursing homes/ skilled nursing facilities/residential aged care facilities. Three studies took place in the emergency department. The rest of the studies occurred in mixed settings, such as combined inpatient/outpatient department studies or nursing homes and community.

### Study populations

All studies reported on interventions targeting adults, with 66% of these in older adults (≥65 years old). 35 studies targeted populations with polypharmacy.

Definitions of polypharmacy varied from ≥5 to ≥15 medications across studies. 20% of studies targeted patients with specific diagnoses, including dementia, chronic disease and psychiatric disorders. One study targeted GPs treating older adults directly and a further study was directed at physicians with high rates of benzodiazepine prescribing to older adults.

### Focus of intervention

120 studies were provider-focused, 38 had a mixed focus towards provider and patient and 18 studies were patient-focused. Of the patient-focused studies, 7 used the EMPOWER brochure (Figure 3).

**Figure 3.**
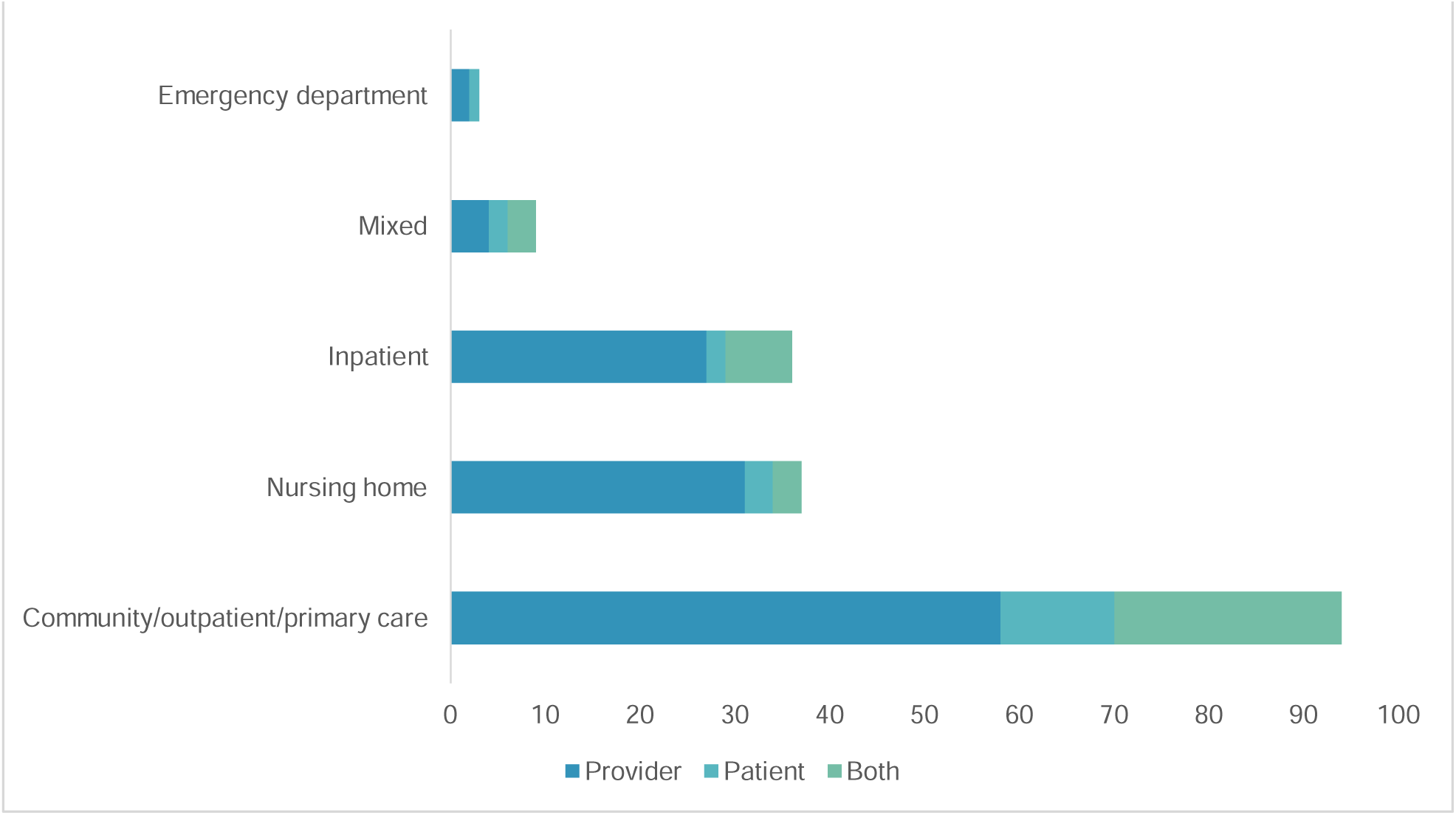
Setting and Focus of Interventions.

**Figure 4.**
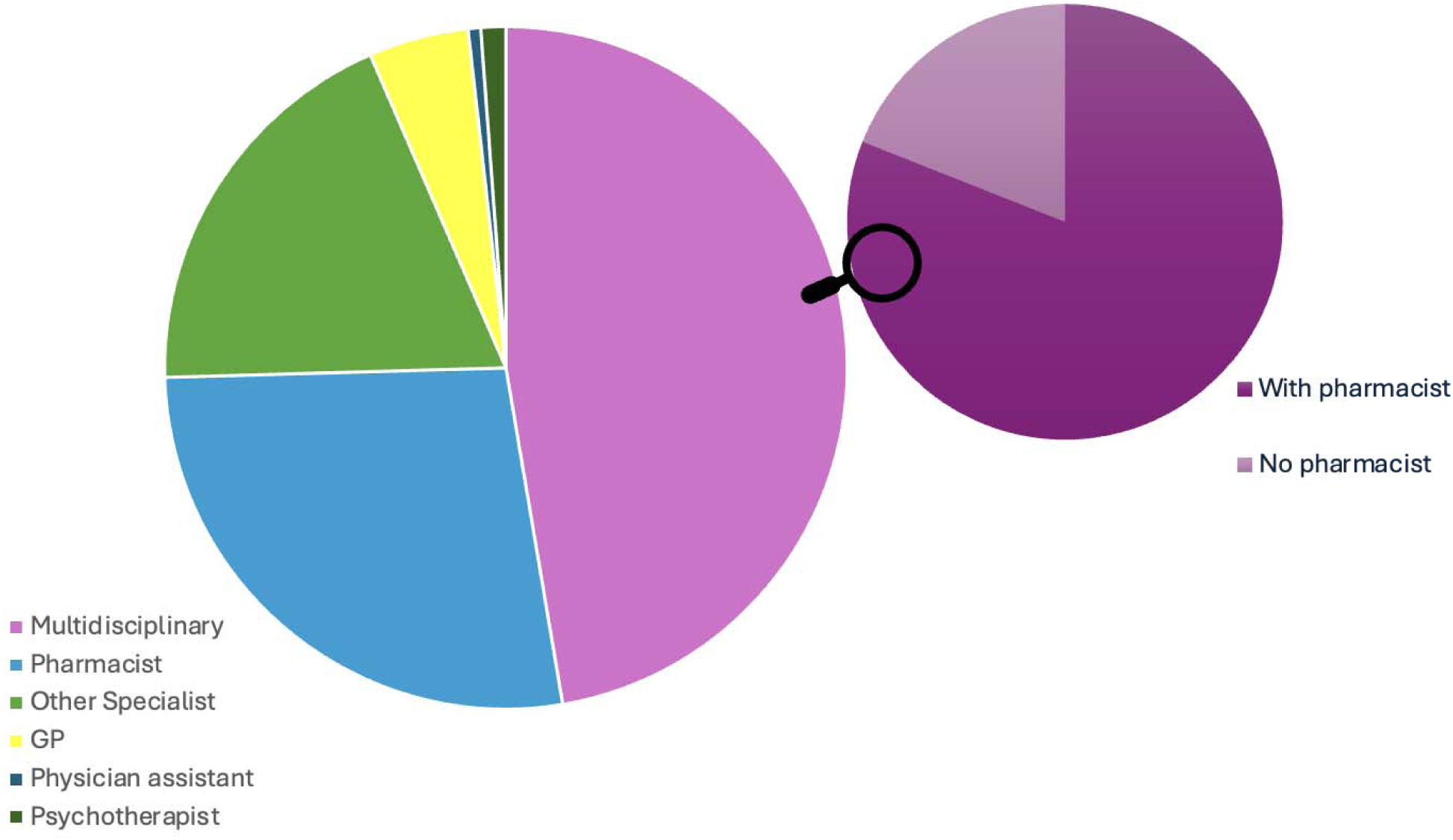
Health professionals involved in deprescribing.

### Professionals involved

In terms of disciplines involved, 45% (n=80) reported on multidisciplinary interventions, involving varying combinations of professionals including pharmacists, GPs, other specialists and other healthcare professions. The vast majority (81%) of the multidisciplinary studies involved pharmacists with other healthcare professionals, of which 64 (80%) included specialist physicians and other healthcare professionals (mainly nurses), and 18 (22%) involved GPs without a pharmacist but other professionals.

In terms of single discipline studies, 46 studies involved pharmacists, 20 interventions involved GPs or primary care physicians only, and 29 studies were carried out by specialists. The most common specialities involved in deprescribing were internal medicine physicians, psychiatrists, clinical pharmacologists and geriatricians.

One study reported on an intervention by physician assistants, and two studies reported on interventions carried out by psychotherapists.

### Medications targeted

51% (n=92) of the studies included had a broad approach without targeting a specific medication. 19 of these specifically mentioned potentially inappropriate medications as identified by Beers or STOPP criteria, three looked at drugs to avoid in the elderly, and three at drugs at high risk of potentially inducing falls. Of studies with specific medication targets, 7% (n=13) looked at multiple medications or classes. The remainder dealt with only one medication or class.

Benzodiazepines or benzodiazepine receptor agonists were addressed in 21 studies. Benzodiazepines were the sole focus of 17 of these studies and appeared in combination with other medications in the remaining papers. 13 studies targeted anticholinergic/sedative medications,18 studied PPI deprescribing, with 14 studies having them as the sole target. Other studies involved psychotropics (n=5), antipsychotics (n=4), antidepressants (n=2), aspirin (n=3), statins (2) and other lipid-lowering agents.

To identify target medications for deprescribing, 50% (n=90) of the studies used specific guidelines. The most frequently used guidelines were Beers and STOPP with 44 and 35 studies, respectively. In many cases, a combination of guidelines was used to identify potential targets. In some cases, these tools were integrated into the Electronic Medical Record (EMR) of the facility.

### Content of interventions

Inspired by Wang et al’s^13^ review on deprescribing in older adults, we identified common broad components of interventions: provider education, patient education, medication reviews, use of clinical decision support systems and behavioural therapy.

53% of studies (94) reported on interventions with an educational component targeting either providers, patients or both.

#### Provider education

Several studies involved provider-directed education before commencement or during implementation of deprescribing interventions. Provider education consisted of in-person teaching, formal tutorials, interactive seminars and/or practice meetings. They focused primarily on cessation principles or best practices for physicians to prescribe for specific populations, such as nursing home residents or patients on antipsychotics. For example, a study assessing a primary care nurse-led intervention in benzodiazepine deprescribing began with two one-hour long seminars on benzodiazepine usage and withdrawal, and training on assessing patients’ need for pharmacological support.^17^

Other strategies used were peer interviews with GPs, ongoing professional development with pharmacists^18^ and multiprofessional in-person workshops with nurses, pharmacists, physicians and managers.^19^ For instance, one study used an educational intervention involving prescription monitoring and periodic meetings by a dedicated “Choosing Wisely” group comprised of two clinicians, a biostatistician, and an expert in quality and patient safety.^20^ Another study used the chronic care model with three training sessions of four hours for participating GPs on narrative-based doctor-patient dialogues.^21^ One study involved peer education to reduce inappropriate prescribing, delivered by GPs who attended pre-study training sessions and visited GP groups twice during the study period.^22^ In one intervention, an educational workshop to reduce inappropriate prescribing for elderly patients involved a multifaceted intervention with educational materials, class-specific algorithms, and two PCP-led half-day educational workshops.^23^

Other studies provided online-only education for providers or only provided educational materials, such as deprescribing guides, information leaflets and online education regarding polypharmacy in older patients, training videos or templates for medication reviews.^24^ Sometimes the educational materials were sent by post.^25^

A few studies had an educational segment involving training with four e-learning modules as well as an on-site workshop.^26,27^ The COSMOS intervention for deprescribing in nursing homes started with an educational component, with a two-day educational seminar (covering communication with relatives and patients, pain assessment and treatment, rationale and method of medication review, role playing and patient centred discussion) for nurses selected to be ambassadors, other nurses, physicians and pharmacists. Ambassadors also received educational material to facilitate training of other nursing home staff.^28^ Providers often received training around various deprescribing tools and guidelines including the Screening Tool of Older Persons’ Prescriptions (STOPP)^29^, the Good Palliative-Geriatric Practice (GPGP) method^30,31^, the Garfinkel method^32^, and the drug burden index (DBI).^33^

#### Patient education

Patient-directed education was mostly via written materials about the importance of being involved in one’s own medications and information sheets for specific drug classes, providing rationale for their withdrawal as well as general advice. Several studies provided patients with the EMPOWER brochure, a patient-empowerment educational booklet. Some were specifically aimed at reducing benzodiazepines,^34–36^ one targeted PPIs and sulphonylureas,^37^ while others targeted inappropriate prescribing in dialysis patients, or more broadly for older people.^38,39^ In some interventions, usually in inpatient setting, patients also received a counselling session.^40^

In a pharmacist-led deprescribing program, patients received face-to-face education on the rationale for deprescribing in a medication management clinic.^41^ An intervention involving an inpatient discharge review service to facilitate deprescribing in over 65-year-olds provided patient-directed education during a discharge interview.^42^ In two studies, chronic pain patients were educated on the benefits of opioid reduction before dose reduction.^43,44^ In one trial, benzodiazepine users received a structured educational interview, which was either followed up in person or with written instructions, with the aim of benzodiazepine deprescribing.^45^

#### Both provider and patient education

Five studies involved educational components for both patients and providers. The D-PRESCRIBE trial used a pharmacist-led intervention to reduce inappropriate prescribing in older patients, providing educational materials for both groups on medication appropriateness, alternatives, and tapering aids.^46^ The OPTIMIZE trial, targeting older adults with cognitive impairment and polypharmacy, offered a two-part educational intervention with brochures and a PATD questionnaire for patients and families, and monthly deprescribing materials for clinicians.^47^ A study on aspirin cessation for primary prevention in adults over 70 provided provider education on updated guidelines and safety, alongside patient education through PowerPoint presentations and flyers.^48^ Gannon et al.’s study focusing on anticholinergic deprescribing in a community mental health setting involved multimodal provider education, with online education modules created by pharmacists and psychiatrists and web streaming videos, patient infographic materials, small group sessions and case consultations regarding anticholinergic medications.^49^

#### Medication review

Many interventions involved an initial review of all medications taken by the patient, which could include only prescribed medications or be extended to over-the-counter medications in some cases. In many studies, this was the first element of a stepwise deprescribing process. Overall, 136 studies (78%) involved a medication review component.

Medication reviews were carried out by pharmacists only in 54% of studies, by GPs or other physicians in 26% of studies and the remaining 20% had multidisciplinary medication reviews, usually involving pharmacists and physicians in conjunction.

Pharmacists alone performed medication reviews in both inpatient and outpatient settings. In inpatient settings this was usually part of deprescribing rounds^50^ or in inpatient deprescribing admission services.^51^ Examples of medications reviews in the outpatient setting included a clinic targeting hospitalisation risks in elderly patients^52^ and in an anticoagulation clinic.^53^

Physician-only medication reviews in inpatient setting were undertaken by clinical pharmacologists^54^ and geriatricians as part of a comprehensive geriatric assessment.^55,56^ In a few studies in primary care setting, medication reviews were carried out by GPs, with two studies involving multiple appointments.^21,24^ In two other studies GPs received instruction from pharmacists before carrying out medication review themselves.^57,58^ Physicians alone carried out medication review in a falls clinic.^59^

Physicians and pharmacists collaborated to perform medication reviews across different settings (inpatient, GP practice, psychogeriatric facility, polypharmacy clinic, nursing home) to achieve deprescription for older adults with polypharmacy.^60–63^ One study also involved a further independent medication review by a nurse.^64^

Medication reviews were carried out as face-to-face reviews^65,66^, during a home visit^67^, by telephone^68^, based on EMR^69–71^ or a combination of an interview and EMR.^72,73^ In the Shed-MEDS intervention, medication history was also obtained through a combination of chart reviews, patient interviews and pharmacy calls.^74^

##### Clinical decision support systems

Many studies used web-based tools or clinical decision support systems for deprescribing, including MEDSAFER, which generates individualized reports based on evidence-based guidelines and tapering instructions^75,76^; TRIM, the Tool to Reduce Inappropriate Medications^77,78^; and Intercheck CPSS, which uses Adverse Drug Reaction scores to identify high-risk patients^79^. Other tools included Arriba PPI for deprescribing potentially inappropriate medications^80^, G-MEDSS for high-risk medications^33^, VIONE, a deprescribing tool from the Veterans Health Association^39^, Prima-eDS for patient recommendations^81^, and Check the Meds, which applies Beers/STOPP and Priscus criteria^82^, along with Optimeds and Pharmanurse online tools.^83^

Two studies reported the use of online tools specifically to check drug interactions, using the Epocrates programme^84^ and the Danish national drug interactions database.^85^ One study involved an intervention based on a mobile app, available to physicians on a tablet, which contained information on PIMs, potential alternatives and deprescribing.^86^

Several studies reported the use of electronic medical records, with many of them using it to identify patients using target medication(s)^87,88^ and to notify caregivers about upcoming visits of patients potentially for deprescribing^89^ and reminders.^90^ Some studies involved computerised alert systems integrated into electronic medical records.^91,92^

Several studies reported the use of algorithms, which could be non-class specific^67,93^ or class specific^23^ with two studies referring specifically to PPI specific algorithms^94,95^ and others to benzodiazepines.^96^

##### Behavioural therapy

Three studies reported on the use of behavioural therapy. One study investigated the effectiveness of cognitive behavioural therapy (CBT) and Therapeutic Interactive Voice Response (TIVR) to reduce analgesia use in participants with chronic pain.^97^ Another study compared the use of CBT plus medication tapering vs tapering alone on deprescribing of benzodiazepines in panic disorder.^98^ Another study assessed the effectiveness of dialectical behavioural therapy to reduce polypharmacy in participants with borderline personality disorder.^99^

##### Medication substitution

Some studies used temporary medication substitution to aid deprescribing, commonly in deprescribing of benzodiazepines (melatonin and pregabalin)^100,101^ and PPIs (histamine 2 antagonists).^102^

### Outcomes measured

The vast majority (90%) of studies looked at medication level outcomes such as rates of deprescribing or the mean number of medications before and after intervention as main outcome measure. In 39% of these this was the only outcome measure described. 87 studies reported on patient level outcomes which included adverse drug events, mortality, hospitalisations and emergency visits and health-related quality of life. No studies reported adverse effects with regard to patient level outcomes though one reported deprescribing related effects in three patients which resolved on re-introduction of the deprescribed medications.^103^ 33 reported on economic outcomes including cost-utility analysis and cost-savings. A third of these reported cost savings, one reported a non-significant mean cost increase^58^ and the remaining studies reported no change. 34 studies reported on process outcomes including acceptability, acceptance of recommendations and feasibility. No studies reported on environmental outcomes of deprescribing interventions.

### GRADE rating and impact

Per the GRADE criteria, 23% of studies received a grading of moderate to high (these were RCTs), 32% low (quasi-experimental and RCTs) and 45% very low (observational and pilot studies). Studies generally receive high GRADE ratings when they are well-designed randomised controlled trials with large sample sizes, low risk of bias, consistent results, and direct evidence; ratings may be downgraded due to factors such as methodological limitations, inconsistency of results, indirectness of evidence, imprecision (i.e. wide confidence intervals), or publication bias.

We extracted data on whether the deprescribing intervention had a significant impact on the outcomes, considering the GRADE ratings. We focused on studies rated as moderate to high GRADE to further examine their intervention characteristics. Overall, 76% of these interventions had a significant impact. Notably, 79% of multidisciplinary interventions reported significant effects, compared to 74% for single-discipline interventions (80% for those involving non-GP physicians, 60% for GPs, and 77% for pharmacists).

All three patient-targeted CBT interventions showed significant impact. Among provider-only interventions, 79% were successful, in particular those using stepwise approaches^34,104^, medication review protocols, and guideline-based clinical decision support systems (e.g., STOPP/Beers criteria^27,56,79^) were consistently associated with successful outcomes. Of the interventions targeting both providers and patients, 50% had a significant impact, especially when combined with patient empowerment tools such as EMPOWER brochures.^36,37^

None of the interventions in our review reported negative impacts on patient-level clinical outcomes.

### Discussion

To our knowledge, this is the first scoping review to comprehensively map the global evidence on deprescribing interventions across populations, medications, healthcare settings, and disciplines. The interventions in our review were largely effective, and none of the interventions reported negative patient-level clinical outcomes. Several studies looked at the economic impact of deprescribing interventions and a third of these suggested economies of different scales.^105,106^ None of the studies reviewed assessed the impact of deprescription on environmental outcomes.

Our review broadens the knowledge base by synthesizing implementation features and effectiveness. This broad perspective is essential to inform evidence-based, system-level deprescribing strategies that go beyond local initiatives or isolated clinical trials.

### Implications for Public Health and Policy

Given the global rise in pharmaceutical consumption, the overuse of potentially inappropriate medications has become a public health challenge with clinical, economic, and environmental implications. Beyond patient safety, medication waste contributes substantially to healthcare-related CO2 emissions and environmental contamination through pharmaceutical runoff.^107^ National and local deprescribing policies should thus be seen not only as clinical quality improvement efforts but also as strategies for environmental health and planetary sustainability.^108^

The evidence in our review is unambiguous: deprescribing offers a safe and effective approach to optimizing medication use. The question is not whether to act, but how best to design and scale these interventions. In this respect, our findings are coherent with recent evaluations of pharmaceutical interventions which emphasize that policy effectiveness depends not only on intervention design, but also on the way it interacts with the specific healthcare context in which it is implemented.^9^

While our review identifies key components of successful deprescribing interventions - such as structured multidisciplinary collaboration, active patient engagement, and use of clinical decision-support tools - the challenge remains in scaling these models, across diverse healthcare settings. This requires nuanced consideration of policy contexts, reimbursement structures, and system-level incentives that influence implementation.

Recent realist evaluations suggest that deprescribing initiatives are more likely to succeed when embedded within explicit national health policies, supported by centralized infrastructure for coordination, and designed with active engagement from stakeholders including both providers and patients.^9^

Successful implementation depends also on healthcare organisation including institutional capacity, financial and behavioural incentives. Effective policy design must account for these factors, as one-size-fits-all approaches are unlikely to succeed. Policymakers need to integrate deprescribing strategies in local public health plans for investing in education, monitoring, and feedback mechanisms to support sustainable behavioural change.

### Future Directions

Despite the breadth of this review, several important gaps in the deprescribing literature remain. Most existing interventions target older adults, with limited attention given to younger adults, working-age populations, or those with chronic non-communicable diseases such as pain disorders or psychiatric conditions. This narrow focus may overlook important opportunities for early intervention and prevention of long-term polypharmacy. Moreover, very few studies explicitly examined how deprescribing interventions perform across diverse patient groups. Further research is needed to assess whether interventions are equally effective across different socioeconomic, ethnic, linguistic, or geographic populations.

As underlined in earlier research^109^, most studies included in our review assessed outcomes over a limited time horizon, often months. Longer-term evaluations are necessary to understand whether deprescribing interventions lead to sustained reductions in medication burden, improved health outcomes, or changes in healthcare utilization. Also, while our review captured many dimensions of the interventions used, we often lacked information on implementation issues and contextual constraints. This limits our understanding of transferability and scalability ^9^ Finally, although the environmental impact of overprescribing is substantial, no studies in our review evaluated environmental outcomes, such as reduction in pharmaceutical waste or carbon footprint. This represents a major missed opportunity for linking deprescribing to broader sustainability goals.^110^

### Limits

Several limitations of this scoping review should be noted: First, by design, we included all interventional studies regardless of their methodological strength or GRADE scoring. This allowed for a more inclusive mapping of real-world deprescribing practices but limits the ability to draw strong conclusions about causality or comparative effectiveness. Second, the included studies varied widely in their populations, settings, interventions, and outcome measures. This precluded meta-analysis or comparisons across intervention types and limited our ability to quantify overall effectiveness. Third, it is likely that studies reporting positive outcomes are overrepresented in the literature, while negative or null findings may remain unpublished.^111^ This “file drawer problem” poses a risk of bias. We cannot rule out the possibility that the evidence base paints an overly optimistic picture. Finally, we did not include grey literature or program evaluations published outside of academic journals. As a result, certain interventions, particularly those in low-visibility settings, may have been missed. Nonetheless, we believe that our approach provides a valuable insight for identifying broad patterns, common components of successful interventions, and directions for future policy-oriented research.

## Conclusion

The overuse of medications is a major threat to both public and environmental health. Our review demonstrates that deprescribing can be achieved safely and effectively across a range of settings, and it should be integrated in collaborative, patient-centred, guideline-based, and context-sensitive approaches. Moving forward, the challenge lies not in proving that deprescribing works, but in embedding it into coherent public health policy, supported by national public health strategies tailored to local realities.

## Supporting information

detailed search strategy

detailed data extraction

## Data Availability

All data produced in the present work are contained in the manuscript

## Notes

### Competing Interest Statement

The authors have declared no competing interest.

### Funding Statement

This study did not receive any funding

