## Supplementary material for "What may work in deprescribing? A scoping review of intervention types, targets and outcomes": detailed search strategy

Search Strategy

**Inital search**

Database: Pubmed

((((policy[Title/Abstract]) OR (policies[Title/Abstract])) OR (intervention[Title/Abstract])) OR (interventions*[Title/Abstract]) AND ((fha[Filter]) AND (english[Filter] OR french[Filter])))

AND ((((("deprescriptions"[MeSH Terms]) OR ("deprescribe"[Title/Abstract])) OR ("deprescribed"[Title/Abstract])) OR ("deprescribing"[Title/Abstract]) AND ((fha[Filter])

AND (english[Filter] OR french[Filter]) AND (2010:2024[pdat]))) AND (polypharmacy[Title/Abstract] AND ((fha[Filter]) AND (english[Filter] OR french[Filter]))) AND ((fha[Filter])

AND (english[Filter] OR french[Filter]))) Filters: Abstract, English, French

Database: Web of Science

(((TS=(policy)) OR KP=(policy)) OR KP=(intervention)) OR KP=(interventions) AND (TS=(deprescription*))

OR KP=(deprescription)

Years 2010-2024

((TS=(polypharmacy)) OR KP=(polypharmacy)) OR AK=(polypharmacy)

AND (((TS=(policy)) OR KP=(policy)) OR KP=(intervention)) OR KP=(interventions)

Years 2010-2024

**Subsequent search**

Database: Pubmed

(inappropriate*[title/abstract]) and ((((polic*[title/abstract]) or (intervention*[title/abstract]) and ((fha[filter]) and (english[filter] or french[filter])))

and (((("deprescriptions"[mesh terms]) or ("deprescribe"[title/abstract])) or ("deprescribed"[title/abstract])) or ("deprescribing"[title/abstract]) and ((fha[filter])

and (english[filter] or french[filter]) and (2010:2024[pdat])))) not ("systematic review"[publication type]))

Database: Web of Science

((TS=(polic*)) OR TS=(intervention*)) AND TS=(inappropriate*)

AND (TS=(deprescription*)) OR TS=(Deprescrib*)

Years 2010-2024
